# Using PARIHS framework to design a community-based COVID-19 intervention in rural Ghana

**DOI:** 10.1101/2021.03.30.21254353

**Authors:** Shadrack Frimpong, Moro Seidu, Sam Kris Hilton

## Abstract

This study utilized the Promoting Action on Research Implementation in Health Services (PARIHS) framework to guide the design of the evidence-based practice – COVID Preparedness & Outbreak Prevention Plan (CoCoPOPP) for rural communities in Ghana. Through a participatory academic-community team discussion, interactive dissemination, review of evidence about community-based interventions during Ebola, HIV/AIDS, and Influenza outbreaks via snowball sampling, continuous discourse within the design team, feedback from other local stakeholders and national experts, the evidence-based intervention was developed consistent with the PARIHS framework. By applying the three core elements of the PARIHS framework (that is, evidence, context and facilitation), the study developed orientation, logistic needs and planning as well as social mobilization. The components also included participants recruitment and training, communication, research and M&E plan, execution and technical assistance and facilitation with three overall aims: (1) meet a pressing health need during the COVID-19 pandemic in local underserved settings, (2) ensure that the strategy is informed by high-quality evidence from similar interventions in past outbreaks and (3) evaluate and learn from research on interventions to garner data for organizational use and to share insights on pandemic management and control with the Ghanaian government, wider global health and education community. Hence, CoCoPOPP can be implemented across other rural communities in Ghana and beyond, particularly in other Sub-Saharan African countries with similar cultural settings.

## 1. Introduction

The novel coronavirus disease has thrown the entire world into disarray – hundreds of thousands of lives have been lost, economies have come to a halt, and the urgency to stop its spread grows daily. As of March 19, 2021, there have been over 123 million confirmed COVID-19 cases worldwide, of which 69.9 million people have recovered and 2.72 million have died (Johns Hopkins, 2021). Leading the pack are the United States of America, Brazil, and India. Accordingly, developing regions such as Africa also continues to experience a surge in infections – the continent has recorded over 4 million confirmed cases (with South Africa being the most drastically affected country), 3.57 million recoveries, and 106,280 have died (Galal, 2021). Ghana, one of the African top 10 nations with a high prevalence of COVID-19, has confirmed 89,682 cases, 85,761 recoveries and 725 deaths as of March 22, 2021 (Ghana Health Service, 2021). Yet these numbers are just the tip of the iceberg given that only 970,048 people out of the country’s 30 million populace have been tested (Ghana Health Service, 2021).

Reported statistics in Ghana and Africa are highly underestimated and do not capture community transmissions, particularly in rural areas (Paintsil, 2020). Rural communities in developing countries including Ghana are considerably more vulnerable to COVID-19 than urban areas (Anant et al., 2020; Jubayer et al., 2020) where the illiteracy rate is high; women-46% and men-28% (Ghana Statistical Service, 2015) and the population is relatively old (Mba, 2010). Mba (2010, 2009, 2007, 2004) ascertained that, rural communities in Sub-Saharan Africa particularly Ghana are ageing faster, as young people move to urban areas and migrants return to their rural communities as they grow old and retire from employment. This makes rural communities even more vulnerable to COVID-19 as the risk of getting severely ill from the virus increases with age (CDC, 2020). Furthermore, comorbidities such as diabetes, cardiovascular diseases, and lung diseases are highest among Ghana’s elderly population (Amoah et al., 2002). Research shows that people with the aforementioned comorbidities are at higher risk for severe illness or death from COVID-19 (CDC, 2020).

Hence, the healthcare challenges posed by the COVID-19 pandemic are even direr in rural areas, where over 70% of the country’s agrarian population already struggle to access healthcare. Yet, very little to no effort has been made by the government to support rural communities. The quality of COVID-19 management support delivered in most rural areas in developing countries including Ghana falls short of that demonstrated to improve health outcomes (Anant et al., 2020; Jubayer et al., 2020; Quaglio et al., 2020; Mitra, 2020). There has neither been an intervention plan nor a comprehensive strategy or design for implementation in Ghanaian rural communities that met their unique cultural and socio-demographic needs. Cocoa360, a non-profit organization in rural Western Ghana that employs a farm-for-impact model to sustainably finance the education of young girls and provide subsidized healthcare, facilitated one of the rural responses to COVID-19. The organization rolled out a collaborative intervention called COVID Preparedness & Outbreak Prevention Plan (CoCoPOPP) in the eight rural communities it currently serves. CoCoPOPP is a concerted effort designed to ensure that rural inhabitants are educated about COVID-19, have access to Personal Protective Equipment (PPE) and high-quality healthcare services through the elimination of treatment fees for Respiratory Tract Infection (RTI) cases.

Oftentimes, public health interventions are not effectively implemented, resulting in unintended negative consequences such as waste of resources. Nonetheless, Dane and Schneider (1998) suggest that five aspects influence the adoption and implementation of an intervention. These include fidelity, dosage (intervention strength), quality, participant responsiveness and program differentiation. Therefore, the gap between the development of effective public health intervention and far-flung sustained adoption in communities is best attained through strategies that consider how the intervention itself can be adopted to meet the needs of the population, structures, and various practitioners (Rabin et al., 2008; Kilbourne et al., 2007). Such strategies should also aim at maintaining the fidelity, or consistent delivery of the components required for the intervention to be effective (Rabin et al., 2008; Kilbourne et al., 2007). However, while there is some literature on the design and implementation of public health interventions, there is limited information in a usable form for public health decision-makers, program planners and practitioners (MacDonald et al., 2016).

Since little is known about how to best design a pandemic response for rural communities in Ghana, Cocoa360 designed a comprehensive intervention titled **‘**Cocoa360’s COVID Preparedness & Outbreak Prevention Plan (CoCoPOPP)’ to reduce the spread of COVID-19 in rural Ghana. The design of CoCoPOPP brought together participatory research and dissemination in a process described as **‘**interactive dissemination**’**. Stakeholders working at different levels in rural communities, research institutions, and the health system were engaged to obtain, analyse and interpret systematic reviews, and essential WHO literature about recent outbreaks in Africa, to identify the impact of community-based interventions that were implemented, identify priority gaps in key areas of health care provision and reflect on the fundamental barriers and enablers as well as to propose strategies for advancement.

In this paper, the Promoting Action on Research Implementation in Health Services (PARIHS) framework was used to analyse the design of the CoCoPOPP intervention to develop an evidence-based intervention for rural Ghanaian communities. The PARIHS framework was conceived by Kitson et al. (1998) to help professionals successfully implement research into practice. Thus, the framework facilitates evidence to use at the implementation level and considers the broader implementation context (Nilsen, 2015; Harvey & Kitson, 2015). The framework consists of three core elements: the level and nature of evidence, the context in which the research is to be applied, and the facilitation of the implementation process (Kitson et al., 1998). This paper aims to describe the design of CoCoPOPP while utilizing the PARIHS framework to enhance its successful integration into other rural communities. Further, we highlight lessons for development professionals in rural areas in a bid to scale up participatory knowledge translation research and to facilitate engagement at a system level.

### 1.1 Overview of CoCoPOPP Intervention

CoCoPOPP is a two-part pilot intervention to address the COVID-19 pandemic in the eight rural communities Cocoa360 serves, namely, Tarkwa-Breman (TB), Techimantia, Doktaso, Bayereagya, Krofofrom, Nkran-Dadieso, Fantekrom and Afukey, all located in the Prestea-Huni Valley district in the Western Region of Ghana. Based on the low literacy levels in the community, and from published evidence about successful Ebola management in rural Liberia; CoCoPOPP implementation began with an intensive community involvement activity coupled with baseline initiatives such as research, recruitment and training of social mobilizers and community health education. The community radio, community leaders and other health service providers within the communities provided support for the intervention, and encouraged residents to participate, by taking the lead in announcing the launch of the intervention, and its goals. Next, the intervention enabled the communities to have access to PPE, high-quality healthcare services through the elimination of treatment fees for RTI cases, subsidies and abatements at our medical facility, while concurrently engaging with the community, conducting research to monitor outcome measures, and supervising our social mobilization team. The subsidies component was in line with Cocoa360’s farm-for-impact model, where user fees (provider consultation, and clinic registration fees) of the cost of healthcare were eliminated with revenues from the most recent cocoa harvest from the TB community farm. The successful implementation of this evidence-based intervention involved the chiefs and elders of the community, and a village committee (VC) which serves as a liaison between the larger TB community, and Cocoa360.

The unique rural and remote locations of the communities present a strong opportunity to conduct research, and gain insights on the pandemic management and control with the Ghanaian government, as well as the wider global health and education community. This would be an extremely crucial resource for the control and management of future pandemic in similar settings. CoCoPOPP, therefore, educated the community about COVID-19, its symptoms, risk factors, preventive and control measures such as sanitation and hygiene, and social distancing. Furthermore, CoCoPOPP strengthened COVID-19’s control capacity in the eight (8) communities by ensuring easy availability of PPE, elimination of treatment fees for RTI cases and subsidizing the cost of healthcare utilization. Finally, CoCoPOPP helped to monitor, evaluate and learn from (conduct research on) intervention to collect data for organizational use, and to share insights on epidemic management and control with the Ghanaian government, wider global health and education community.

## 2. Methods

Developing a comprehensive intervention especially for rural communities is more challenging than simply combining abstract from existing theories. This study utilized an evidence-based decision-making framework, PARIHS, to inform the design of the CoCoPOPP intervention to reduce the spread of COVID-19 in TB – a rural community in Ghana. TB community and the surrounding villages access primary health care through community-controlled and government-managed services specifically designed to meet their needs as well as through private general practices.

The study took advantage of academic-community-based partnerships between community non-profit organization (Cocoa360), Ghana Health Service (GHS)-Prestea Huni Valley Health Directorate (PHVHD), TB Community Clinic (TBCC) and TB Community-Based Health Planning and Services (TBCHPS), University of Ghana, Yale University, Vanderbilt University and the rural communities Cocoa360 serves. The key organization that worked with community leaders to co-design the intervention is cocoa360. Cocoa360 worked closely with the VC, which is the primary decision-making board consisting of respected and elected members from Cocoa360’s partner communities. The VC ensures that the organization’s operations are aligned with the needs and cultural dynamics of the communities they serve.

### 2.1 Study Framework

The study applied PARIHS framework based on its focus on the implementation stage of intervention to design CoCoPOPP which is a culturally sensitive and context-dependent intervention. With a strong emphasis on evidence, context and facilitation, PARIHS framework provides significant guidelines for ensuring that interventions when implemented, achieve the highest positive outcomes, with minimal unintended negative consequences. Several empirical studies have provided support for the PARIHS framework by demonstrating that successful implementation is a function of evidence, context and facilitation (Kitson et al., 2008; Cummings et al., 2007; Estabrooks et al., 2007; Ellis et al., 2005; Wallin et al., 2005; Kitson et al., 1998).

PARIHS framework was developed to describe implementation success in health care settings (Kitson et al. 1998). This framework states that successful implementation is a function of three elements: evidence, context and facilitation. Each element and its respective sub-elements are rated on a continuum from low to high. Kitson et al. (1998) establish that the most successful implementation occurs when: the evidence is scientifically robust and matches professional consensus and target population needs (‘high’ evidence); the context is receptive to change with sympathetic cultures, strong leadership, and appropriate monitoring and feedback systems (‘high’ context); and there is appropriate facilitation of change with input from skilled external and internal facilitators (‘high’ facilitation)(Rycroft-Malone et al. 2002). Thus, this study specifically applied the evidence, context and facilitation elements of PARIHS framework in designing CoCoPOPP intervention for successful implementation to reduce the spread of COVID-19 in vulnerable rural populations in the eight rural communities that Cocoa360 operates in.

### 2.2 CoCoPOPP Intervention design

In March 2020, the intervention design team, which consisted of the executive and research team of Cocoa360, the VC, community leaders, a physician assistant, and health educators at TBCC, a coordinator at GHS and a university-based research team engaged in a participatory process to design CoCoPOPP in community primary care settings, serving vulnerable rural populations. Guided by the PARIHS framework, the process involved a review of evidence about community-based interventions during Ebola, HIV/AIDS, and Influenza outbreaks, continuous discourse within the design team, as well as feedback from other local stakeholders and national experts.

### 2.3 Research Evidence Generation

A snowball sampling approach was performed to obtain systematic reviews, and essential WHO literature about recent outbreaks in Africa, and the impact of community-based interventions that were implemented. The searches and reviews were limited to articles published between 2014 and 2020. The articles were selected through titles and abstracts while considering recent outbreaks in Africa.

### 2.4 Inclusion Criteria

The selection of articles for review was based on inclusion criteria. The articles had to report on the results of research and contain information about disease outbreaks, precautionary and protective measures adopted, and the impact of interventions and means of adoption. The articles had to be published between 2014 and 2020; report on vulnerable communities or countries in Africa. Moreover, the articles had to report on an epidemic or pandemic outbreak. All types of health research were considered, that is, both quantitative and qualitative research, as well as literature reviews and a few published essays by WHO.

### 2.5 Exclusion Criteria

Articles in summary and anecdotal form were not included in the literature search and review. Articles on health education were excluded. GRADE-CERQual (Confidence in the Evidence from Reviews of Qualitative Research) was the main tool used to assess the quality of evidence since most included manuscripts utilized qualitative methods (Lewin, 2018). Hence all articles that were rated not ‘high confidence’ based on the CERQual assessment were excluded.

### 2.6 Analysis

The articles were analysed by the GRADE-CERQual approach developed by the GRADE (Grading of Recommendations Assessment, Development and Evaluation) team. The articles were assessed by how much confidence placed in the findings of the qualitative evidence syntheses by considering the methodological limitations, relevance, coherence, and adequacy of data. Four levels were applied to describe the overall assessment of confidence: high, moderate, low or very low.

## 3. Results

### 3.1 CoCoPOPP Intervention based on PARIHS framework

The study designed the intervention by applying the evidence, context and facilitation elements of the PARIHS framework. These elements interact in robust and complex ways to influence CoCoPOPP implementation effectiveness. Comparing the fundamental components of CoCoPOPP to the framework of PARIHS, it was observed that the components scored high ratings in terms of the construct: – evidence (research, professional experience, and community preference) context (culture, leadership and evaluation) and facilitation (characteristics, role and style of the facilitators) [see Table 3].

### 3.2 Satisfying Evidence: Evidence Generation and Quality Assessment

The intervention was scientifically robust. It relied on the research of published sources, matches professional opinion reached by the group as a whole and meets the need of the TB and its surrounding communities because it depends on the community opinion and routine information derived from the community, without valuing only one of its forms to the detriment of others.

### 3.3 Scientific Robustness of CoCoPOPP

Out of the 39 published articles that were considered for the design, a total of 19 articles were selected for inclusion. However, 11 articles were purposely reviewed and analysed because they showed minor concerns with high confidence in their methodological limitations, relevance, coherence, and adequacy of data based on the GRADE-CERQual approach. Conversely, the remaining 8 were not considered in the design. The design was based on research articles from disease outbreak from 2014-2020 [see Table 1]. Eleven major articles were reviewed on Ebola, HIV and influenza outbreaks which provided useful information on disease outbreaks, precautionary and protective measures of disease outbreaks, the means and impact of the adoption of intervention in rural communities for the design team. The articles considered have minor concerns on their methodological limitations, relevance, coherence, and adequacy of data. Similarly, most of the articles have high CERQual assessment of confidence.

**Table 1:**
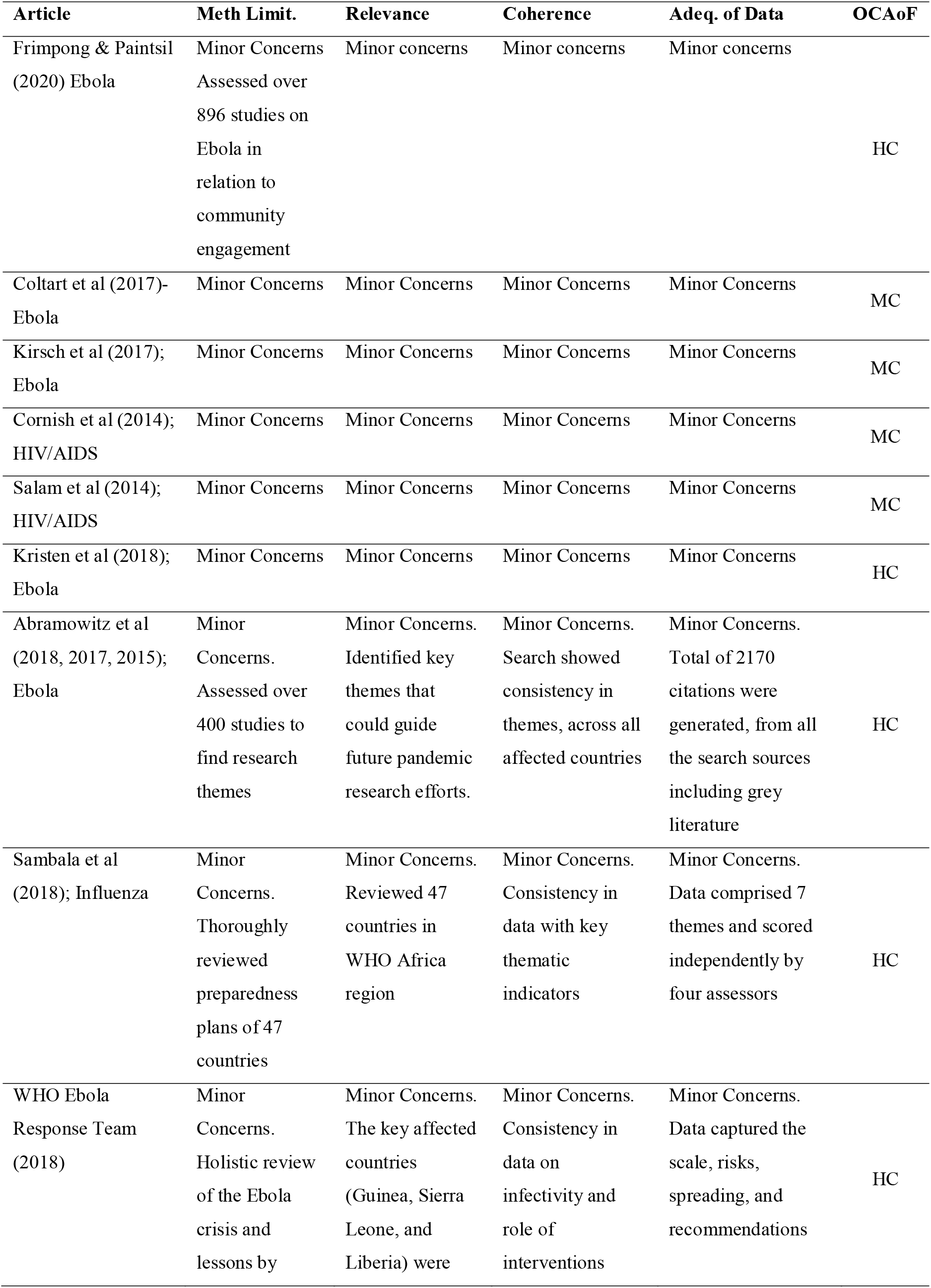
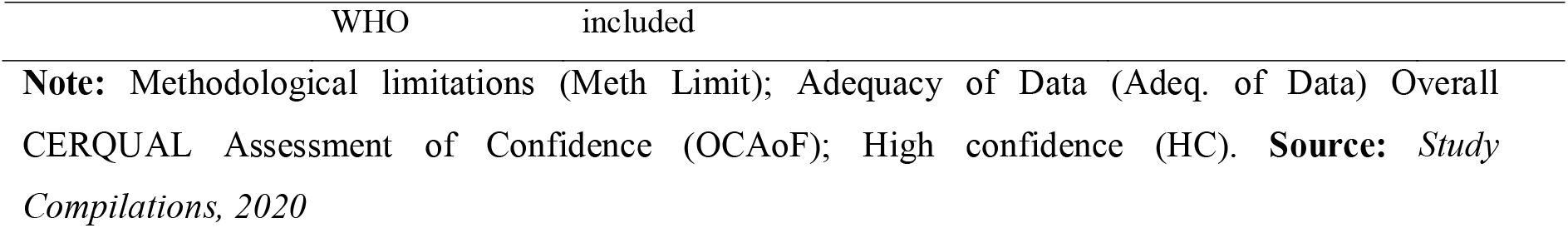
List of articles on epidemic analysed for CoCoPOPP.

### 3.4 Evidence from Clinical Experience

The study also relied on the opinion and experiences of professionals. Physicians and clinical practitioners from GHS-PHVHD, TBCC and TBCHPS who understand the socio-cultural dynamics, disease prevalence, demographics and health care needs and services utilization of TB were part of the design team. They were able to share their consensus opinion through the participatory effort of the design process, and in all stages of the design, they build a strong consensus towards the intervention.

### 3.5 Evidence-based on Community Experiences

The design process ensured that the intervention met the preference and needs of the community; hence members of Cocoa360 within TB were included in the design team. The design process was led by the VC and community leaders, an indication of complete involvement of the community in the design process and the effort to ensure that the outcome of the initiative addresses the community-specific problem. Representatives of the eight communities, the VC were deeply involved in the planning and design of CoCoPOPP. The VC and Cocoa360 shared and analysed information and made recommendations that were relevant to the local practical context. CoCoPOPP’s design was driven by evidence given that information derived from research, clinical experience and local practical context were respectively from robust methodology (such as RCT), consensus and met community needs [see Table 3].

### 3.6 Satisfying Context in the design of CoCoPOPP

The design of evidence-based CoCoPOPP was highly sensitive to the needs of the target population. The intervention was design to be aware of the culture of the communities while considering the leadership, monitoring and feedback systems in the rural communities they were done in.

#### 3.6.1 Culture Context of CoCoPOPP

The intervention was designed to meet the cultural fit of the TB community and the surrounding villages. As part of the implementation strategy of the intervention; it was specified that,

> “…*CoCoPOPP is first presented to the Chief and elders of TB for feedback, support and suggestions. Also request that a Community Leader (preferably the local Chief) announce CoCoPOPP to the community, highlighting the community’s risk, and the intervention’s potential impact, and encouraging interested residents to sign up for social mobilization roles…*” (Cocoa360, 2020)

The implementation strategy gave a greater mandate to the chief and elders (who are the custodians of the communities and villages), to approve of the intervention before it was unveiled or implemented. Hence, the following was documented in the design of the implementation strategy:

> “…*After approval from community leaders and Cocoa360’s Village Committee (VC), we shall secure necessary logistics…*” (Cocoa360, 2020)

The intervention was designed to ensure that the community leads and champions the communication aspect of the intervention.

> “…*Request Community Leaders to Champion CoCoPOPP: Take the lead on telling the community about CoCoPOPP and cultivating their support*…” (Cocoa360, 2020)

Moreover, the design of the intervention-implementation strategy also ensured that the community members did not only benefit from the intervention but also took active roles in the implementation process and were treated as experts (*see excerpts from the intervention document below*).

> *“…Requesting community leaders (preferably the local Chief and VC) to encourage interested residents to sign up for social mobilization roles…” and*
>
> *“*…*All participants recruited for the surveys and focus group discussion are treated as experts”* (Cocoa360, 2020)

The study ensured that all participants were respected and treated as experts, reimbursed their travelling costs (if any), gave participants gift (such as prepaid phone cards after the interview /focus discussion) or gifts that might be useful for the participant in the context of the cultural norms of the community instead of cash. The issue of acceptability, trust, recognition, and respect was minimized by engaging the community leaders and VC in introducing CoCoPOPP to the communities. Moreover, the recruitment announcement of CoCoPOPP was first delivered by local leaders at a community meeting. Similarly, community leaders were included in the discussions to promote community members’ participation.

CoCoPOPP was also designed to promote learning; research the intervention to collect data to try new and different techniques, for organizational use, and to share insights on epidemic management and control with the Ghanaian government, wider global health and education community (*see excerpts from the intervention document below*).

> *“…the intervention presents a strong opportunity to conduct research, and gain insights on epidemic management and control with the Ghanaian government, as well as the wider global health and education community. This will be an extremely crucial resource for the control and management of future epidemics in similar settings…”* (Cocoa360, 2020)

All these implementation measures guaranteed that CoCoPOPP was effective in minimizing the spread of COVID-19 in the community while following the cultural dynamics of the people.

#### 3.6.2 Leadership Context of CoCoPOPP

CoCoPOPP was designed to ascertain clear roles and objectives among the stakeholders [see Table 2] involved in the intervention. The stakeholders within each group work together as a team and share power. For instance, TBCC healthcare workers work closely with each other and have broad authority in treating their clients. Each of the micro teams was coordinated by the Cocoa360 managers to ensure harmony and good communication among the teams. Because of the clearly defined roles, responsibilities, objectives and effective coordination specified for each of the stakeholders and among the various team units, CoCoPOPP enrollment was characterized by a high sense of leadership [see Table 3].

**Table 2:** Stakeholders involved in the implementation of CoCoPOPP.

| Primary stakeholders | Secondary Stakeholders |
| --- | --- |
| Community leaders (Chief and elders, Village Committee) | University of Ghana |
| Community members | Ghana Ministry of Health (MoH) |
| Cocoa360 executives and directors | Vanderbilt University |
| TBCC healthcare workers | Yale University |
| Cocoa360 Research Team | Donors |
| Social mobilizers |  |
| community liaison |  |
| information flow manager |  |
| Data collectors |  |
| TBCHPS |  |

**Source:** *Compiled from CoCoPOPP implementation document, 2020*

**Table 3:** CoCoPOPP satisfying PARIHS framework elements and sub-elements.

| Elements | Sub-element | Rating |
| --- | --- | --- |
| **Evidence** | Research | High |
|  | Professional (Clinical) Experience | Moderate |
|  | Community preference | High |
| **Context** | Leadership | High |
|  | Culture | High |
|  | Evaluation (Measurement) | High |
| **Facilitation** | Characteristics of facilitator | High |
|  | Role of facilitator | High |
|  | Style of facilitator | High |

**Source:** *Compiled from CoCoPOPP implementation document, 2020*

### 3.7 Evaluation of CoCoPOPP

Evaluation is one of the key fulcrums CoCoPOPP leverages on; where the intervention strategy allows that interdisciplinary investigators (such as the University of Ghana, Yale University, Vanderbilt University, MoH, GHS and Cocoa360) took part in the monitoring and evaluation of the intervention. Below is an excerpt from the initiative, elaborating how CoCoPOPP was consistent with the PARIHS framework’s sub-element evaluation.

> *“…A strong team of interdisciplinary investigators at the University of Ghana, and Yale University in partnership with the Ghana Ministry of Health (MoH), Ghana Health Service (GHS), Cocoa360 and Village Committee shall research to monitor and evaluate the CoCoPOPP intervention…”* (Cocoa360, 2020)

The intervention package further allows for data collection before, throughout and at the end line, to measure the effectiveness of all possible activities and outcomes. Likewise, the design of the evidence-based intervention also factored in all the necessary metrics to estimate the possible individual and team performance, activities, outputs, outcomes and impact of the intervention. CoCoPOPP also emphasized feedback on individual, team, and the intervention performance on the community;

> *“…Consistent with our community engagement principles as an organization, we will continue to update VC and community chiefs and elders about progress* → *materials distributed; cases being seen…”* (Cocoa360, 2020)

The robustness and consistency of evaluation (that is, the presence of routine monitoring systems) throughout the phases of CoCoPOPP can be described as high based on the concept of the PARIHS framework [see Table 3].

### 3.8 Satisfying Facilitation in the design of CoCoPOPP

Facilitation is a sub-element in the PARIHS framework, which is a function of implementation success and also influential in overcoming the barriers to evidence-based practice (Rycroft-Malone et al., 2013). The designers of CoCoPOPP took into account good facilitation in the design process by soliciting inputs from relevant internal and external facilitators.

#### 3.8.1 Characteristics of the facilitators

The intervention was designed to factor inputs from skilled internal facilitators such as community leaders (chief and elders, VC), Cocoa360 executives, TBCC healthcare workers, social mobilizers, Cocoa360’s research team, and data collectors while also utilizing inputs from adept external facilitators from the University of Ghana, Ghana MoH, Vanderbilt University and Yale University. These facilitators exhibit characteristics consistent with that of opinion leaders, change agents, champions, educational outreach workers, and linking agents in the implementation strategy to promote high facilitation.

The chief and elders within the facilitation of CoCoPOPP are the opinion leaders from the local community, viewed by the community as highly credible, respected sources of influence (via authority, status and representativeness), ability to persuade and have a broader span of influence across the communities. The VC within the facilitation of CoCoPOPP are the champions who are internal to the community and implementation organization, have visionary qualities and overwhelming enthusiasm in the intervention. Moreover, the Cocoa360 executives, TBCC healthcare workers, and Cocoa360’s research team, social mobilizers and data collectors are the internal change agents that promoted the action of CoCoPOPP. These facilitators have strong interpersonal and communication skills; are knowledgeable and understanding; and have earned the trust and respect of the community because of their consistent interaction with the people for at least two years. The external facilitators of CoCoPOPP are educational outreach workers; they are topic experts that are external to the intervention setting and knowledgeable about their area of specialization. They met with other facilitators to provide useful information about the evidence-based intervention and provide feedback as and when necessary.

#### 3.8.2 Role and style of the facilitators

These skilled facilitators had clearly defined roles to achieve a specific objective in the practice of CoCoPOPP and to ensure consistency in the delivery process. Facilitators especially those directly involved in enrolment of the practice have experience of at least two years in the environment of the intervention area and fully aware of the possible challenges they are likely to face, hence are flexible and show empathy when dealing with the people and tenacious in overcoming obstacles. Thus, the planners of CoCoPOPP considered high facilitation [see Table 3] of change with input from adept internal and external facilitators who have high respect, credibility, empathy, authority and influence with clearly defined roles, flexibility and consistency.

## 4. Discussion

The PARIHS provided a pragmatic framework for rendering guidance toward the design and development of an evidence-based strategy to successfully influence the implementation of a COVID-19 Preparedness & Outbreak Prevention Plan intervention in a rural setting. The consideration of PARIHS framework in the design process pointed out various barriers and facilitators of implementation that may not have otherwise been explicitly accosted. The inclusion of the VC, chief and elders of TB in the design team, recruiting participants from the community as social mobilizers and data collectors, and Cocoa360 leading the development of the intervention boosted community engagement, acceptance, participation and ease facilitation of CoCoPOPP practice. The importance of the considerations are consistent with the assertions of Blumenthal (2006); Bringle and Hatcher (2002) that the thoughtfulness of considerations lead to easy access to community members homes, families and residence, identify other stakeholders critical to the effective adoption of the intervention that was not considered earlier, the provision of local leadership and guidance on the most appropriate means of setting the intervention and promote community enthusiasm and involvement in data gathering.

There have been many interventions just like CoCoPOPP in the past which failed to meet implementation or adoption success partly due to the inability of the implementation to meet the needs, attitudes and beliefs of the community members, cultural and social context. Thus the PARIHS framework was developed by Kitson *et al*. (1998) to meet implementation success in health care organizations. CoCoPOPP was therefore designed with thorough guidance from PARIHS framework by simultaneously considering the evidence, context and facilitation as the key pivot of the design instead of a hierarchy or linearity of cause and effect of the elements to influence implementation success in a rural setting. From Table 3 it was observed that the design of CoCoPOPP was scientifically racy and corresponds to professional consensus and TB and its surrounding communities need (‘high’ evidence). The context was open to change with harmonic cultures, strong leadership, and refined evaluation, monitoring and feedback systems (‘high’ context). There was also appropriate facilitation from competent and experienced internal and external facilitators (‘high’ facilitation). The design of CoCoPOPP was, therefore, consistent with the guidance proposed by (Rycroft-Malone, 2007; Rycroft-Malone, 2004; Rycroft-Malone et al., 2002; Kitson et al., 1998).

The context, in which CoCoPOPP was implemented, was crucial to the success of implementation. The context of CoCoPOPP was ranked high because the study ensured that the evidence is practised in an environment with strong leadership, strong awareness of community and embedding organizational culture and high monitoring, evaluation and feedback systems. CoCoPOPP, therefore, considered all the following context factors in its design; participant-mix, staff-mix, access to resources/equipment, community culture, implementation organizational climate, financial disincentives, an academic affiliation of Cocoa360, other healthcare affiliation with TBCC, evaluation, provision of education, information and communication flow, research activities, stress, community readiness for change, uncertainty, uncontrolled events, support, participants turnover, leadership, decision-making structure, and autonomy. These contextual factors are to effectively promote the success of the implementation of evidence to practice (Bostrom et al., 2009; Yano, 2008; Francke et al., 2008; Koh et al., 2008; Scott et al., 2008; Dijkstra et al., 2006; Meijers et al., 2006). Therefore, the design process considered broad contextual factors to significantly minimize intervention planning and roll-out.

Another key feature of CoCoPOPP is its high facilitation designed to emphasize the purpose and role of facilitation with the skills and attributes of the facilitators (both internal and external). CoCoPOPP depended on the experiences and skills of opinion leaders, change agents, educational outreach workers, and linking agents, where the purpose of facilitation was categorically defined to achieve specific goals and the development of processes to enable effective teamwork. The characteristics of CoCoPOPP facilitation which is also affected by the skills and attributes of the facilitators corroborates with the finding of Harvey et al. (2002) that there is a facilitation continuum that ran from task-specific to holistic.

## 5. Conclusion

The PARIHS framework was employed to design the CoCoPOPP intervention to reduce the spread of COVID-19 in Ghanaian rural communities. By applying the three core elements of the framework and its related sub-elements, such as evidence (research, professional experience, and community preference) context (culture, leadership and evaluation) and facilitation (characteristics, role and style of the facilitators), the designers developed orientation, logistic needs and planning, social mobilization, participants recruitment and training, communication, research and M&E plan, execution and technical assistance and facilitation components of the intervention with three overall aims: (a) meet a pressing health need during the COVID-19 pandemic in a rural setting:-TB and its surrounding villages via awareness and education about COVID-19, promotion of access to PPE, access to free treatment of RTI cases and provision of subsidies and abatements at community based health facility – TBCC; (b) ensure that the intervention strategy is informed by robust, high-quality evidence from similar interventions in past outbreaks and (c) monitor, evaluate and learn from research on interventions to garner data for organizational use, and to share insights on pandemic management and control with the Ghanaian government, wider global health and education community. Hence, CoCoPOPP can be implemented across other rural communities in Ghana and beyond, particularly in other Sub-Saharan African counties with similar cultural setting.

## Data Availability

Data is available upon request.

## Acknowledgments

We gratefully acknowledge the thoughtful comments we received on earlier drafts from Priya Bhirgo, Julian Addo, and Irma Lee. Their comments and dialogue helped strengthen the paper. Thanks also to Newlove Gershon Nkebe for the useful discussion of the ideas presented in this paper. In addi-tion, the insightful depth reflected in comments provided by Cocoa360 team members also considerably strengthened the paper.

